# Extraction-free protocol combining Proteinase K and heat inactivation for detection of SARS-CoV-2 by RT-qPCR

**DOI:** 10.1101/2020.12.16.20248350

**Authors:** Valeria Genoud, Martin Stortz, Ariel Waisman, Bruno G. Berardino, Paula Verneri, Virginia Dansey, Melina Salvatori, Federico Remes Lenicov, Valeria Levi

**Author notes:** Corresponding authors (VG), (VL). These authors contributed equally to this work (Valeria Genoud, Martin Stortz).

## Abstract

Real-time reverse transcription PCR (RT-qPCR) is the gold-standard technique for severe acute respiratory syndrome coronavirus 2 (SARS-CoV-2) detection in nasopharyngeal swabs specimens. The analysis by RT-qPCR usually requires a previous extraction step to obtain the purified viral RNA. Unfortunately, RNA extraction constitutes a bottleneck for early detection in many countries since it is expensive, time-consuming and depends on the availability of commercial kits. Here, we describe an extraction-free protocol for SARS-CoV-2 detection by RT-qPCR from nasopharyngeal swab clinical samples in saline solution. The method includes a treatment with proteinase K followed by heat inactivation (PK+HID method). We demonstrate that PK+HID improves the RT-qPCR performance in comparison to the heat-inactivation procedure. Moreover, we show that this extraction-free protocol can be combined with a variety of multiplexing RT-qPCR kits. The method combined with a multiplexing detection kit targeting N and ORF1ab viral genes showed a sensitivity of 0.99 and a specificity of 0.99 from the analysis of 106 positive and 106 negative clinical samples. In conclusion, PK+HID is a robust, fast and inexpensive procedure for extraction-free RT-qPCR determinations of SARS-CoV-2.

## Introduction

The outbreak of the coronavirus disease 2019 (COVID-19), caused by SARS-CoV-2, was declared a pandemic by World Health Organization on March 11, 2020. Confronted by the lack of a vaccine or specific treatment, early detection, isolation and contact tracing are fundamental strategies for reducing the spread of this disease [1].

RT-qPCR is the gold standard method for SARS-CoV-2 determinations [2]. These assays typically involve collecting the clinical specimen usually with nasopharyngeal swabs, extracting the RNA from the sample and analyzing the presence of the viral RNA through RT-qPCR.

Nowadays, the RNA extraction step is considered a bottleneck of SARS-CoV-2 RT-qPCR determinations [3]. This procedure can be automatized but commercial extraction robots are very expensive and may not be affordable for most clinical labs in low-income countries. In contrast, manual RNA-extraction is a quite cumbersome procedure that includes several washing and centrifuging steps at a Biosafety level 2 laboratory and relatively expensive extraction kits. In Argentina, a trained lab technician processes approximately 24 samples every 1.5 hours. Additionally, the skyrocketing increment on the clinical use of RT-qPCR due to the COVID-19 pandemic led to a worldwide shortage of RNA purification reagents [4, 5] thus, many research groups around the world have concentrated their efforts on either simplifying or eliminating the extraction step [6-16].

Björn Reinius at the Karolinska Institute in Sweden [17] optimized an extraction-free sample preparation protocol that only includes a very simple heat-inactivation step at 95°C during 5 min (HID method). Through HID RT-qPCR, they were able to detect with high sensitivity the relatively short N1 and N2 regions of the viral RNA which are preserved after the heating step. This group also performed an exhaustive analysis of media formulation compatible with the HID method [17]. More recently, the Centers for Disease Control and Prevention (CDC) has approved a similar thermal-inactivation protocol (1 min at 95°C) that can be used with Quantabio UltraPlex 1-Step ToughMix (4X) and is sufficient for viral inactivation [18]. This protocol also detects N1 and N2 sequences of the viral RNA.

Many protocols traditionally used for extraction-free preparation of clinical samples included treatments with proteinase-K (PK), a commonly used protease that degrade RNases and hence, it helps to preserve RNA integrity[19, 20]. A recent work suggested that PK may facilitate extraction-free determinations of SARS-CoV-2 although a major loss in sensitivity with a ∼6-units shift in CT values on the determination of the envelope (E) gene [21] may prevent its uses in clinical analysis. Other recent preprint [22] tested a protocol based on PK and heat inactivation using a reduced number of samples (N=17) in UTM medium with relative low CT values, and claim a good sensitivity on the determination of the E gene. Although these results might seem contradictory with recent findings showing suboptimal detection of the E amplicon on HID samples in UTM medium[17], the efficiency of the PCR is very dependent on the detection kit, transport medium brand, thermocycler, etc.

Here, we combined these strategies and optimized the PK+HID method, a very simple protocol for extraction-free RT-qPCR determinations of SARS-CoV-2. Our results position PK+HID as an inexpensive, versatile and reliable alternative to bypass the traditional RNA extraction procedure and suggest that it could substantially contribute to improve the testing capacity in countries with limited resources and facilities.

## Materials and Methods

### Sample collection

Nasopharyngeal swabs were collected during screening of non-hospitalized symptomatic cases and close contacts to these cases at diverse hospital and clinical centers at the Buenos Aires area (AMBA, Área Metropolitana de Buenos Aires), Argentina. Swabs were deposited in 2-5 ml of saline solution. These clinical samples were processed at the Instituto de Investigaciones Biomédicas en Retrovirus y SIDA (INBIRS, Buenos Aires, Argentina), a specialized center currently dedicated to SARS-CoV-2 diagnosis by RT-qPCR. For routine diagnosis, RNA extraction was performed from 300 µl of swab samples using a Chemagic 360-D (Perkin-Elmer), an automated extraction equipment, according to the manufacturer’s instructions. This procedure does not include an RNA-concentration step.

This study used the remaining volume of anonymized samples that had been collected for clinical diagnosis. Under these circumstances, and for studies involving development of COVID-19 diagnostic tools, our IRB (the Research Ethics Committee from Fundación Huésped in Buenos Aires, Argentina) deemed unnecessary to obtain informed consent from the patients.

### PK+HID sample preparation

Nasopharyngeal swabs deposited in 2-5 ml of saline solution were maintained up to 24 h at 4°C. PK+HID samples were prepared 10-24 h after RNA extraction unless indicated otherwise. 10 µl of a solution 10 mg/ml Proteinase K (Promega, Madison, WI, USA or Inbio Highway, Tandil, Argentina in FlashPrep solution of Inbio Highway) were placed in 0.2 ml PCR tubes. Then, 90 µl of the nasopharyngeal swabs samples previously vortexed were transferred to each of these PCR tubes. Please, notice that the PK solution should be added to the tubes before transferring the samples and not the other way around. This procedure minimizes the work-hazard since tubes with samples are not reopen until inactivated. It also provides flexibility in the workload of clinical labs because the preparation of PCR tubes with PK solution could be done outside the Biosafety Level 2 facility in a clean, nucleic acid-free area.

For experiments with variable PK concentrations, 10 µl of PK stock solutions with the adequate concentration were added. Samples were incubated at 55°C for 15 min and then at 98°C for 5 min in a thermal cycler with heated lid. Finally, inactivated samples were cooled to 4°C and kept at this temperature until RT-qPCR analysis.

### RT-qPCR

One-step reverse transcription and qPCR of N1 and N2 viral regions were performed using the GoTaq® Probe 1-Step RT-qPCR System (Promega, Madison, WI, USA) and the 2019-nCoV CDC Diagnostic Panel (Integrated DNA Technologies, Coralville, IA, USA) according to manufacturer’s instructions. The N1 and N2 viral regions and the human RP (internal control) were detected in singleplex 20 µl-volume reactions using 5 µl of purified RNA or 5 µl of PK+HID samples unless specifically stated in the text. Thermal cycling steps were: 45°C for 15 min, 95°C for 2 min, and 45 cycles of 95°C for 3 s and 55°C for 30 s. RT-qPCR was performed on a StepOnePlus Real-Time PCR System (Applied Biosystems).

One-step reverse transcription and qPCR of N and ORF1ab viral genes were performed following routine diagnostics procedures at INBIRS using the DisCoVery SARS-CoV-2 RT-PCR Detection Kit (Safecare Biotech Hangzhou, China) according to manufacturer’s instructions. The N and ORF1ab viral regions and the human RP internal control were detected in multiplex reactions. Thermal cycling steps were: 50°C for 5 min, 95°C for 30 s, and 45 cycles of 95°C for 5 s and 60°C for 34 s. RT-qPCR was performed on a CFX96 Real-Time System.

One-step reverse transcription and qPCR of N, E and RdRp viral genes were performed following routine diagnostics procedures at INBIRS using the GeneFinderTM COVID-19 Plus RealAmp Kit (Gene Finder, Korea) according to manufacturer’s instructions. The N, E, RdRp viral regions and the human RP internal control were detected in multiplex reactions. Thermal cycling steps were: 50°C for 20 min, 95°C for 5 min, and 45 cycles of 95°C for 15 s and 58°C for 1 min. RT-qPCR was performed on a CFX96 Real-Time System (BioRad).

### Infectivity assays

Vero E6 cells were grown in Dulbecco’s modified Eagle’s medium (DMEM) supplemented with 10% fetal bovine serum, 1% penicillin/ streptomycin, 0.5 mg/ml Amphotericin B and 1% L-glutamine (all from Gibco). Cells were seeded on a 24-well plate at 3.5 × 10^5^ cells/ml one day prior to infection. Nasopharyngeal swab samples were diluted 1:2 in DMEM, filtered through a 0.22 mm-pore-filter and added to cell cultures for 1 h. Then, cultures were washed and incubated for 3 days. When the cultures showed cytopathic effects, the supernatant was collected for detection of SARS-CoV-2 by RT-qPCR, and the culture was terminated. In the absence of cytopathic effects, the supernatant was passed to a new culture. When no cytopathic effect was observed after the 3rd passage, the supernatant was collected for detection of SARS-CoV-2 by RT-qPCR. All infection experiments were conducted in a biosafety level 3 laboratory.

### Loop-mediated isothermal amplification

Reverse transcription, isothermal amplification of the viral genes ORF1a, ORF1b, N and E, and colorimetric detection were performed with COVID-19 Neokit Tecnoami (Neokit S.A.S., Buenos Aires, Argentina), according to manufacturer’s instructions. Briefly, reactions were performed in 0.2 ml PCR tubes mixing 35 µl of reagent and 8 µl of the nasopharyngeal samples. The reactions were run in a thermal cycler with heated lid at 64°C for 1 h. Diagnostics was made according to final color of the solution (violet, negative; blue, positive, S1 Fig).

### Data analysis

qPCR amplification curves obtained in those experiments using DisCoVery and GeneFinder detection kits were analyzed with the CFX Manager software (BioRad) to obtain CT values. Data obtained with 2019-nCoV CDC were analyzed with the free software LinRegPCR [23] to obtain CT values. This software was also used to calculate PCR efficiencies and the initial amount of template copies (in arbitrary units).

## Results

### The treatment of nasopharyngeal swab samples with proteinase-K (PK) improves the performance of the heat inactivation protocol

We used a reduced number of SARS-CoV-2 positive (N=6) and negative (N=3) nasopharyngeal swab samples collected in saline solution to test if their preincubation with PK before the thermal inactivation step improves the performance of the HID extraction-free method. Although previous results [17] suggested that saline solution is suboptimal for direct RT-qPCR detection, this is one of the transport medium recommended by CDC and it is also commonly used to collect clinical samples in Argentina.

In these experiments, we mixed up 90 μl of the samples with 10 μl of PK 10 mg/ml (final concentration 1 mg/ml) in 0.2 ml PCR tubes and incubated them in a thermocycler during 15 min at 55°C followed by 5 min at 98°C (PK+HID samples). As controls, we used heat-inactivated aliquots omitting the preincubation with PK at 55°C (HID samples) and RNA samples obtained through a standard extraction protocol (purified RNA samples) from the same nasopharyngeal swabs. These samples were analyzed by RT-qPCR using the 2019-nCoV CDC Diagnostic Panel which detects the N1 and N2 viral regions and the human RNase P (RP) gene as internal control.

Fig 1a shows that all positive and negative samples were identified using either HID or PK+HID treatments. As examples, Fig 1b shows representative amplification curves obtained for a positive PK+HID sample illustrating that the complex matrix of PK+HID samples did not affect the expected sigmoidal-like shape of the curves. Additionally, the pretreatment with PK consistently resulted in lower CT values than those obtained with HID samples and closer to those determined with purified RNA samples (Fig 1a and S1 Table).

**Fig 1.**
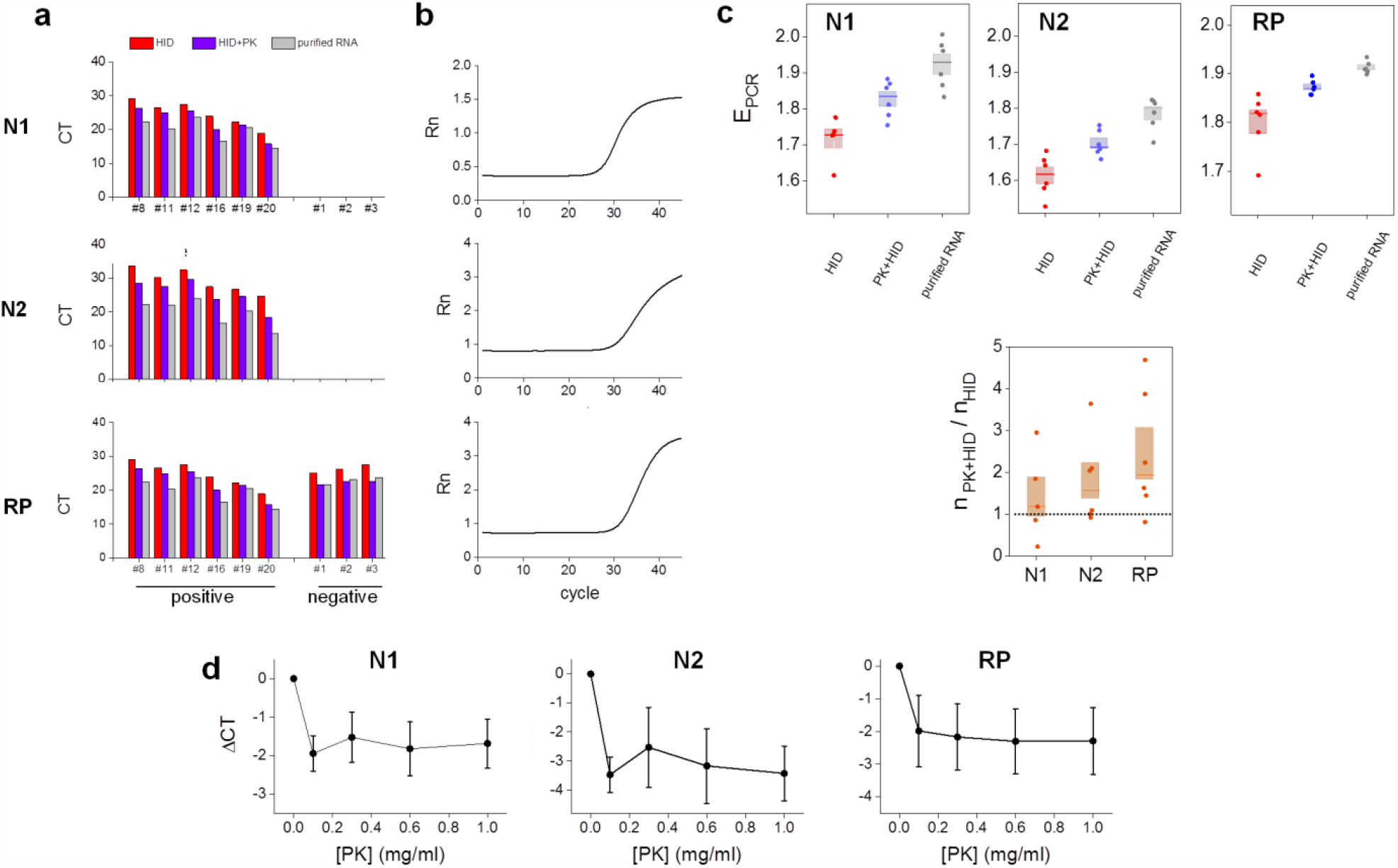
Proteinase K improves the performance of the heat inactivation method in RT-qPCR determinations of SARS-CoV-2. **(a-b)** Positive (#8, #11, #12, #16, #19, #20) and negative (#1, #2, #3) nasopharyngeal swab samples were processed by heat inactivation (HID, 98°C for 5 min); proteinase K treatment followed by heat inactivation (PK+HID, 55°C for 15 min and 98°C for 5 min) or subjected to RNA extraction (purified RNA). The viral N1 and N2 genes and the human RNase P gene (RP) were amplified and detected by RT-qPCR. **(a)** CT values obtained from RT-qPCR analysis of the same samples prepared by the three different methods. **(b)** Representative amplification curves for each gene obtained for one of the positive samples. **(c)** Amplification efficiencies (E_PCR_) and initial amount of amplicon copies (n) in PK+HID samples relative to the corresponding HID samples. The median of each measurement is represented with a line in the bars and the lengths of these bars represent the standard error. **(d)** Positive nasopharyngeal swab samples were subjected to treatment with different concentrations of proteinase K (PK) followed by heat inactivation (55°C for 15 min and 98°C for 5 min). The viral N1 and N2 genes and the human RP gene were amplified and detected by RT- mean difference between CT values obtained in each analyzed condition and the corresponding to HID samples. Mean ± SEM values are represented (N = 5).

The analysis of the amplification curves shows that the PCR efficiency (E_PCR_) was the lowest for the HID treatment followed by PK+HID and then by the RNA extraction treatment (Fig 1c). This result suggests that the PK treatment may contribute to degrade unknown components of the sample matrix that interfere with RT-qPCR. Additionally, we observed a higher PCR amplification efficiency for the N1 viral gene compared to N2, in line with previous results [17, 24]. We also analyzed the amount of amplicon copies in PK+HID relative to that determined in HID samples and observed that this ratio was higher than 1 for N2 and RP suggesting that PK treatment also protected these RNA regions from RNase’s action (Fig 1d).

PK+HID involves the addition of the PK buffer and thus a 10 % dilution of the saline medium used to collect nasopharyngeal swabs. Therefore, we next evaluate if the improvement of PK+HID treatment compared to heat-inactivation is only due to the dilution of the saline medium. We analyzed five positive samples processed by the standard PK+HID protocol or by HID but adding to the samples the same amount of PK buffer without PK (HID’ samples). S2 Fig shows that CT values for PK+HID samples were lower compared to HID’ samples; the analyses of amplification curves indicate that n_PK+HD_ was higher than n_HID’_ supporting that PK contributes with the preservation of the integrity of the viral RNA. Therefore, the dilution of saline solution could not completely explain the better performance of PK+HID. Nevertheless, the efficiency of PCR amplification for HID’ samples was slightly lower only for N2 amplicon suggesting that the dilution of saline solution in PK+HID samples also contributes to increase the efficiency of PCR.

We next tested if using a higher volume of PK+HID samples could improve SARS-CoV-2 detection in the RT-qPCR determinations. Although the sensitivity of the assay is expected to be higher with more copies of the viral RNA template, a previous report also suggested that increasing too much this volume may have the opposite effect due to RT-qPCR inhibitors in the sample matrix [17]. We observed that the amplification curves shifted to lower cycles when using instead of-qPCR determination resulting in lower CT values for the N1 region (0.3 to 1 units, S2 Table). This result suggests that a higher sample volume could contribute to the detection of SARS-CoV-2 in samples with low viral titers.

Finally, we tested if the performance of the PK+HID method depends on PK concentration in the range of 0.1 to 1 mg/ml. Fig 1d shows that CT values obtained for the viral and human amplicons did not change within this PK concentration range. These values were also lower than those observed in the absence of PK, confirming the improving role of PK in this extraction-free method.

### A comparative RT-qPCR analysis with purified RNA samples shows an efficient detection of N1 SARS-CoV-2 region in PK+HID samples

We next ran RT-qPCR determinations of SARS-CoV-2 in PK+HID and purified RNA samples prepared from positive and negative nasopharyngeal swab samples. Based on the results showed in the previous section, we used 1 mg/ml PK for the PK- (i.e. between the volumes assayed in the previous section) in the following RT-qPCR assays. For this particular experiment, we only analyzed the N1 viral gene and the RP human internal control to maximize the number of samples loaded in the PCR multiwell plate.

In these experiments, we correctly identified all positive (27/27) and negative (6/6) samples. Fig 2 also shows that the CT values determined for the viral sequence N1 in PK+HID positive samples strongly correlated with those values measured in purified RNA samples supporting that the PK+HID treatment robustly preserves the information on the viral loads in the swab specimens.

**Fig 2.**
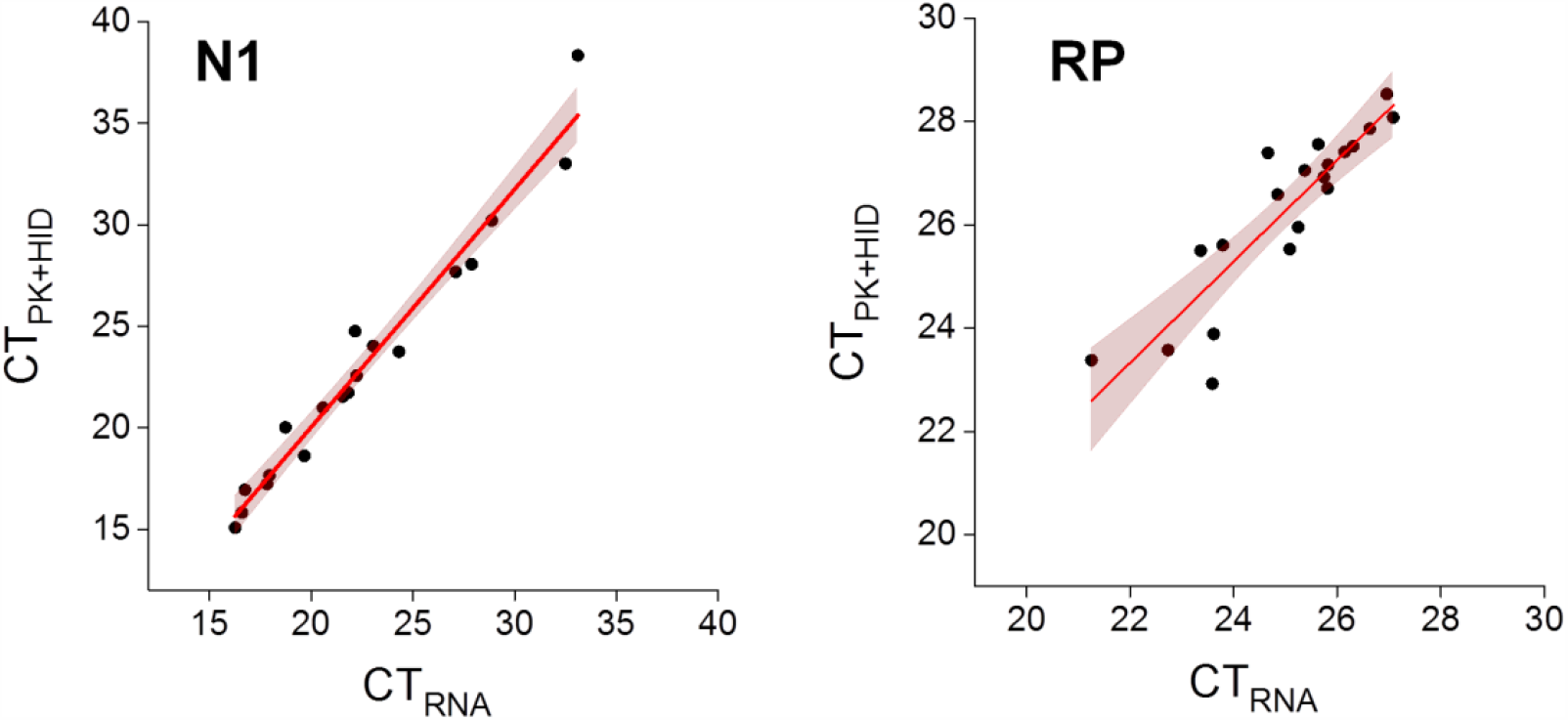
PK+HID method exhibits a similar performance than RNA extraction in RT-qPCR determinations of the SARS-CoV-2 N1 gene. Positive nasopharyngeal swab samples were processed for RNA extraction (purified RNA) or subjected to treatment with PK 1 mg/ml (55°C for 15 min) followed by heat inactivation at 98°C for 5 min (PK+HID). The viral N1 gene and the human RP gene were amplified and detected by RT-qPCR. CT values obtained for the same samples prepared by the two different methods (N = 27), the line obtained by regression of the data (continuous lines) and 95 % confidence bands (pink) are represented. The parameter values obtained from the fitting were: slope = 1.17 ± 0.05 and intercept = −3.4 ± 1.2 (N1 amplicon); slope = 1.0 ± 0.1 and intercept = 1.7 ± 3.0 (RP amplicon).

### The PK+HID method can be used in combination with different multiplexing RT-qPCR detection kits allowing the detection of SARS-CoV-2 with high sensitivity and specificity

In the previous sections, we showed that the PK+HID method resulted in an efficient alternative to bypass RNA extraction when detecting the N1 and N2 regions of the viral RNA. However, most laboratories use PCR kits that target other viral genes often combined in multiplex setups. Therefore, we next evaluated if the improved PK+HID method could be used with multiplexing detections kits.

We first used DisCoVery (Safecare Biotech Hangzhou), a SARS-CoV-2 multiplex RT-qPCR detection kit that targets the SARS-CoV-2 genes N and ORF1ab and human RNase P gene. Relevantly, many worldwide providers offer detection kits targeting one or both of these genes.

Fig 3 shows that the PK+HID treatment of the samples allows the detection of viral and internal control genes. In the case of viral amplicons, the CT values obtained with PK+HID samples linearly correlated with those obtained with purified RNA samples. The slopes for both N and ORF1ab were ∼1 and the y-intercepts were ∼0 suggesting that these sequences are detected equally well in samples prepared with the PK+HID method or through RNA extraction. This observation is also valid for samples with relatively high CT values (>35) indicating that weak positive samples are still detected with the PK+HID method.

**Fig 3.**
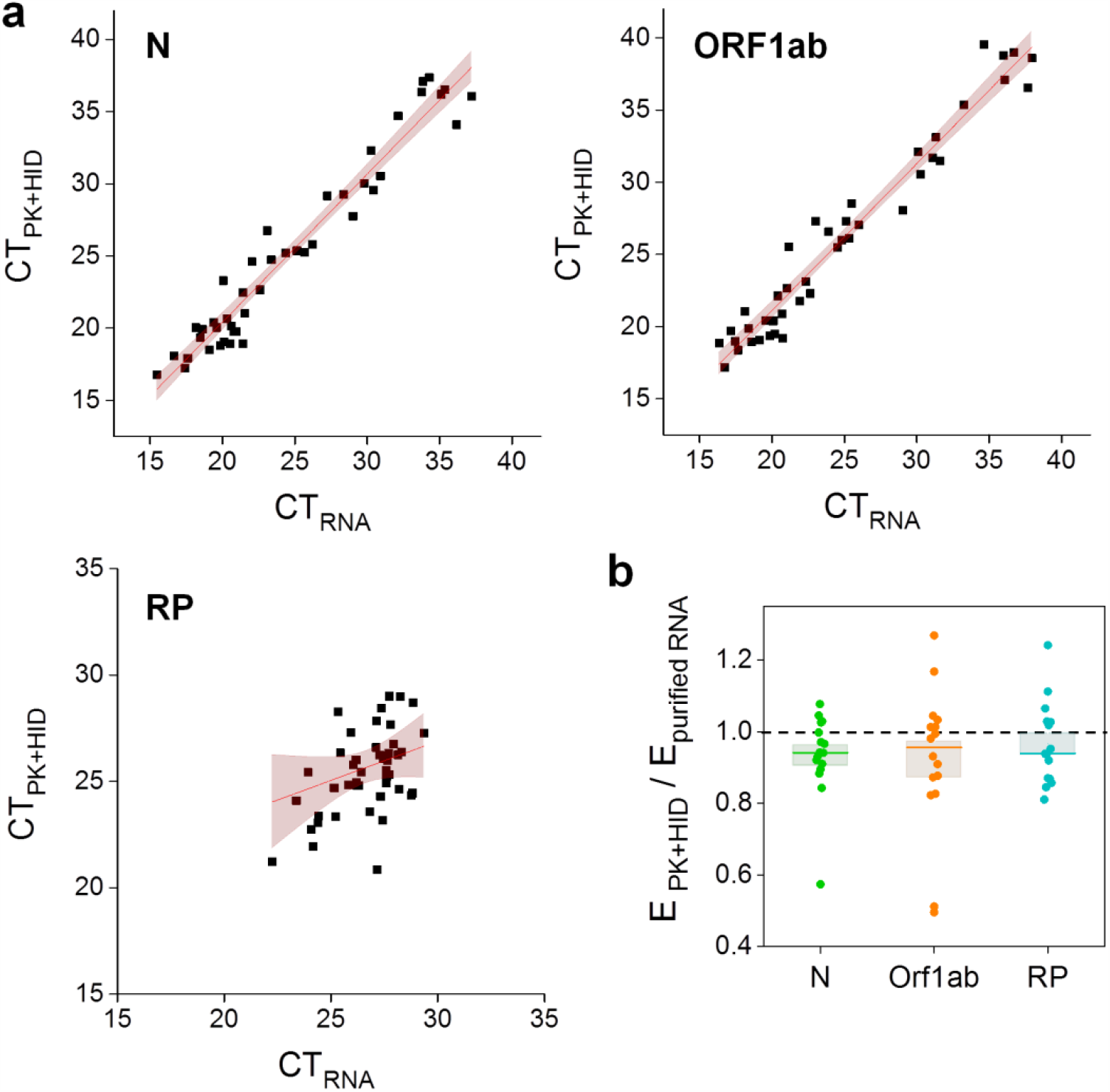
PK+HID method exhibits a similar performance than RNA extraction in RT-qPCR determinations of the SARS-CoV-2 N and ORF1ab genes. Positive nasopharyngeal swab samples were processed for RNA extraction (purified RNA) or subjected to treatment with PK 1 mg/ml (55°C for 15 min) followed by heat-inactivation at 98°C for 5 min (PK+HID). The viral N and ORF1ab genes and the human RP gene were amplified and detected by RT-qPCR. **(a)** CT values obtained for the same samples prepared by the two different methods, the line obtained by regression of the data (continuous lines) and 95 % confidence bands (pink) are represented. The parameter values obtained from the fitting were: slope = 1.02 ± 0.04 and intercept = 0.1 ± 0.9 (N amplicon); slope = 1.02 ± 0.04 and intercept = 0.8 ± 0.9 (ORF1ab amplicon) and slope = 0.4 ± 0.2 and intercept = 15 ± 6 (RP amplicon). Notice that CT values determined for RP spans a smaller range. **(b)** Amplification efficiencies obtained in PK+HID samples (E_PK+HID_) relative to the corresponding purified RNA samples (E_purified RNA_). The mean values obtained for E_purified RNA_ were: 1.98 ± 0.03 (N), 2.02 ± 0.06 (ORF1ab) and 1.94 ± 0.03 (RP). The median of each measurement is represented with a line in the bars and the lengths of these bars represent the standard error (N = 16).

We correctly identified 44/46 positive and 3/3 negative samples. S2 Table compiles the CT values of the false-negative samples (see Discussion). Additionally, the internal control (IC) of two other PK+HID samples was not detected making mandatory repeating the assay; these invalid samples were not included in S3 Table. We determined the amplification efficiency of the viral regions N and ORF1ab and the human gene RP and found that these values were similar for PK+HID and purified RNA samples (Fig 3b). We also compared the amplification curves obtained using this detection kit and swab samples that were either treated by HID or PK+HID or subjected to the RNA extraction procedure (S3 Fig). PK treatment lowered the CT values in comparison to those obtained with HID only, with a mean + SEM difference in CT values of 2.7 + 0.7 (N), 3.1 + 1.0 (ORF1ab) and 1.4 + 0.2 (RP). These results, obtained with a different detection kit, agree with those showed in Fig 1 also supporting that the PK treatment improves the performance of the heat-inactivation method.

The work of Reinius and colleagues [17] confirmed the virus inactivation after HID treatment. As a control, we also run infectivity assays as described in Materials and Methods using 4 nasopharyingeal swab samples with relatively high viral loads (CT values 22.11; 21.32; 22.74 and 19.18 for N amplicon). After the 3rd passage of cell culture, we did not verify any cytopathic effect and the viral RNA was not detected by RT-qPCR showing that the PK+HID treatment also inactivates the virus.

We also analyzed if PK+HID samples can be preserved at 4°C since this could be a common practice in clinical labs. We processed nasopharyngeal samples using the PK+HID treatment and run RT-qPCR determinations after different incubation times at 4°C (0, 20 and 40 h). S4 Fig shows that the CT values for N and ORF1ab genes did not change during the first 20 h and increased at the longest incubation time suggesting that PK+HID samples can be conserved at 4°C for up to 20 h.

A common practice in many clinical labs involves freezing/thawing reagents several times. Therefore, we tested if repeated cycles of freezing/thawing of the PK stock solution may affect the performance of PK+HID. To address this issue, we processed 10 positive samples by PK+HID using a PK solution that was previously subjected to 10 cycles of freezing/thawing or a PK solution thawed only once. The mean differences in CT values between these samples were 0.17 + 0.15 (N), −0.05 + 0.20 (ORF1ab) and 0.01 + 0.14 (RP) (N_data_ = 10) showing that the PK stock solution is stable for up to 10 cycles of freezing/thawing.

To analyze if the PK+HID method can be combined with other multiplexing kits, we prepared PK+HD and purified RNA samples from 94 randomly-selected clinical specimens and ran RT-qPCR determinations using the GeneFinder detection kit which amplifies N, E and RdRp regions of the viral RNA and the human RP gene as internal control.

The analysis of CT values measured in PK+HID and RNA samples showed stronger correlations for the N amplicon in comparison to E and RdRp amplicons (Fig 4a). Additionally, some of the N-positive RNA samples (also considered SARS-CoV-2 positive in clinical diagnosis [24]) showed negative results for the E and RdRp amplicons (3/38 and 4/38, respectively); this difference increased in PK+HID samples with 8 more samples showing negative results for both amplicons while being N-positive. As mentioned above, other authors also reported a poor detection and large right-shifts in CT values for E amplicon in extraction-free samples [17].

**Fig 4.**
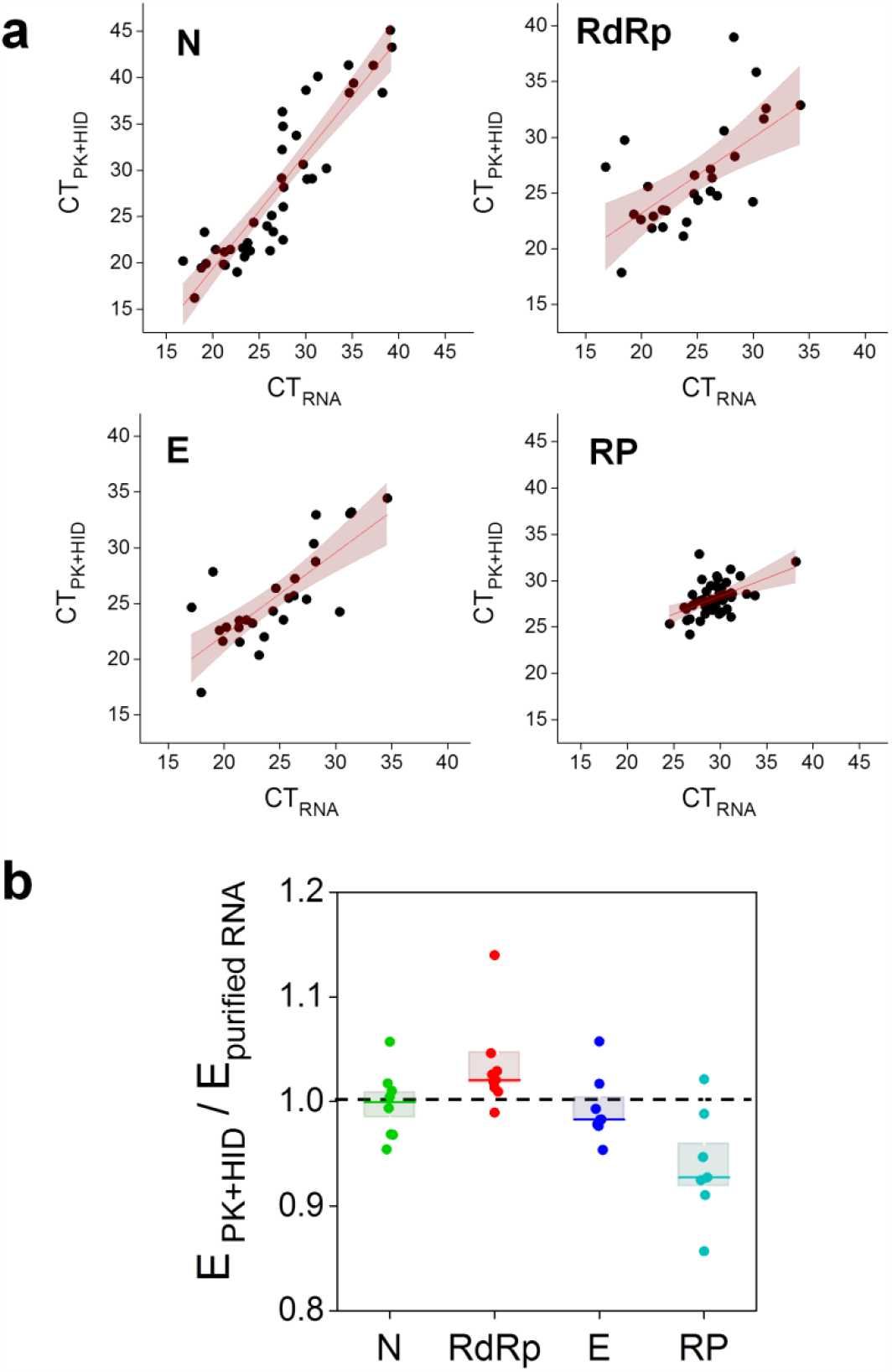
Performance of PK+HID method in RT-qPCR determinations of the SARS-CoV-2 N, RdRp and E genes in randomly-selected clinical specimens. Nasopharyngeal swab samples were processed for RNA extraction (purified RNA) or subjected to treatment with PK 1 mg/ml (55°C for 15 min) followed by heat-inactivation at 98°C for 5 min (PK+HID). The viral N, E and, RdRp genes and the human RP gene were amplified and detected by RT-qPCR. **(a)** CT values obtained for the same samples prepared by the two different methods, the line obtained by regression of the data (continuous lines) and 95 % confidence bands (pink) are represented. RP plot also includes the data obtained in SARS-CoV-2 negative samples and spans a smaller CT range. The parameter values obtained from the fitting were: slope = 1.2 ± 0.1 and intercept = −5 ± 3 (N amplicon); slope = 0.7 ± 0.2 and intercept = 9 ±4 (RdRp amplicon); slope = 0.7 ± 0.1 and intercept = 7 ± 3 (E amplicon) and slope = 0.4 ± 0.1 and intercept = 16 ± 3 (RP amplicon). **(b)** Amplification efficiencies obtained in PK+HID samples (E_PK+HID_) relative to the corresponding purified RNA samples (E_purified RNA_). The mean values obtained for E_purified RNA_ were: 1.94 ± 0.02 (N), 1.97 ± 0.02 (RdRp), 1.93 ± 0.03 (E) and 1.90 ± 0.03 (RP). The median of each measurement is represented with a line in the bars and the lengths of these bars represent the standard error (N = 8).

We also observed that the amplification efficiency of viral amplicons N, E and RdRp was similar for PK+HID and purified RNA samples whereas E_PCR_ of the internal control RP was slightly lower for PK+HID samples (Fig 4b). These results support that the PK+HID treatment contributes to inactivate inhibitors of the PCR reaction.

To quantitatively analyze the performance of the PK+HID treatment combined with the use of GeneFinder kit, we estimated the accuracy, sensitivity and specificity of detection as previously described[25] obtaining values of 0.95, 0.92 ± 0.05 and 0.96 ± 0.04, respectively.

In the validation assay described above, swab samples were first analyzed by the usual RT-qPCR procedure including the RNA extraction step; 10 to 24 h later, we processed the same nasopharyngeal swabs by PK+HID. Therefore, we asked if the extra-time that swab samples remain at 4 C could lead to a partial degradation of viral RNA and consequently, to an apparently lower sensitivity of the PK+HID method. To test this hypothesis, we performed the validation experiment running simultaneously the RNA extraction and PK-HID procedures using 106 positive and 106 negative randomly-selected samples and analyzed the RNA and PK+HID samples by RT-qPCR with the detection kit DisCoVery (S5 Fig); unfortunately, we could not run these experiments with the GeneFinder detection kit used above because the provider run out of stock during the pandemic outbreak. The analyses of the data showed that the accuracy was 0,990 and the sensitivity and specificity were both 0.99 ± 0.01. These results strongly support that PK+HID is a reliable protocol for extraction-free RT-qPCR SARS-CoV-2 determinations.

In this last validation assay, we also analyzed the amplification curves of positive samples with relatively low viral loads (CT values >30) to estimate the relative amount of viral amplicon copies in the PK+HID and RNA samples volumes used in the RT-qPCR determination. S5 Fig shows that the relative amount of viral N and ORF1ab amplicons were in the same order in PK+HID vs RNA samples; particularly, the mean ratio n_PK+HID_/n_RNA_ was 0.7 ± 0.2 (N) and 1.7 ± 0.6 (ORF1ab). This result suggests a similar performance of PK+HID or RNA extraction combined with this detection kit.

Finally, we asked whether PK+HID could be combined with SARS-CoV-2 loop-mediated isothermal amplification kits since they are now widely used in many countries because their simplicity and associated lower costs. S1 Fig includes some examples of positive and negative results obtained with this kit. We could detect 29/29 and 5/6 positive and negative samples, respectively suggesting that PK+HID could be combined with these type of detection kits.

## Discussion

The gold standard method for detection of SARS-CoV-2 in nasopharyngeal swab samples is RT-qPCR, a technique with exquisite sensitivity and high specificity commonly applied to analyze purified RNA samples.

The RNA extraction step has nowadays become the bottleneck in COVID-19 testing, especially during pandemic outbreaks when personnel, resources and biosafety compatible facilities to perform the testing are scarce.

The fast evolution of COVID-19 pandemic and the necessity of performing massive and rapid RT-qPCR determinations to test, trace and isolate SARS-CoV-2 positive patients encouraged the scientific community to evaluate alternative methods to bypass the laborious RNA extraction step. We mentioned above heat inactivation methods that make use of a fast thermal-treatment of the samples and allow the identification of SARS-CoV-2 positive samples targeting N1 and N2 viral genes [17]. These results are consistent with other works that also tested the performance of heat-inactivated respiratory samples in RT-qPCR assays [8, 11, 26-29].

Here, we combined heat inactivation with a pretreatment with PK, protease routinely used in nucleic acid preparations and thus widely available in most clinical laboratories. We mentioned previous works using PK to treat nasopharyngeal swab samples; the protease is also starting to be used in the analysis of saliva samples for the detection of SARS-CoV-2 in extraction-free RT-qPCR determinations [21, 22, 30, 31].

In line with these reports, our data shows that preincubation with PK improves the performance of the heat-inactivation protocol in RT-qPCR assays targeting N1 and N2 viral genes with the 2019- nCoV CDC Diagnostic Panel. However, a validation with a larger number of samples should be done to compare the sensitivities of both methods. We verified that the performance of the PK+HID does not depend on PK concentration in the range of 0.1 – 1 mg/ml; this robustness of the PK+HID method is appealing since small variations in the PK concentration will not significantly affect the results of SARS-CoV-2 determinations and also allows reducing even more the costs of sample preparation for RT-qPCR assays.

Using the 2019-nCoV CDC Diagnostic Panel, we could correctly identify the SARS-CoV-2 positive and negative analyzed samples and, for the positive samples, we obtained CT values in PK+HID samples very close to those of purified RNAs (2.7 units for N1) whereas heat inactivation without PK treatment produces higher shifts in CTs (5 units for N1).

Remarkably, we demonstrated that the PK+HID method could be used in combination with other RT-qPCR kits that target different viral genes in multiplex RT-qPCR reactions. Specifically, we used DisCoVery (targeting N and ORF1ab genes) and Genefinder (targeting N, E and RdRp genes). The detection of these genes was unexpected since the HID method did not provide satisfactory results in the detection of larger amplicons [17]. We found that CT values of both N and ORF1ab viral amplicons detected with DisCoVery were similar to those measured for the purified RNA samples, in line with our observations with the CDC kit.

Although the detection of the E and RdRp genes with the Genefinder kit was weaker than the detection of the N gene, the relatively high detection sensitivity of the PK+HID method estimated from the analysis of 94 randomly selected samples suggests that the PK+HID method can be used as an alternative, extraction-free RT-qPCR analysis of nasopharyngeal swab samples in clinical diagnostics.

S3 Table compiles the CT values of all false-negative samples collected during the course of our work. Most of these samples show high CT values suggesting that they correspond to swab samples with relatively low viral loads.

We should emphasize that the success of an extraction-free method strongly depends on the correct combination of transport medium and detection kit. Merindol et al [32] found that saline solution swab medium impaired the performance of the AllplexTM 2019-nCoV Assay. Although, in our hands, saline solution swab medium could be used with 2019-nCoV CDC, GeneFinder™ COVID-19 Plus RealAmp and DisCoVery SARS-CoV-2RT-PCR detection kits. Since the extraction-free protocol may reduce the efficiency of the PCR, it is critical to select a PCR kit that preserves sensitivity. While most bibliography, as well as our own findings point to N as the most sensitive viral region, ORF1ab gene also showed a good performance with our extraction-free protocol.

## Conclusions

Our results indicate that the combination of proteinase K preincubation with heat inactivation is a very robust and reliable procedure for extraction-free RT-qPCR determinations of SARS-CoV-2. We should mention that preparation of the 94 PK+HID samples required to complete the 96-well PCR plate (2 wells are needed for the positive and negative controls) takes approximately 60 min by one operator whereas manual extraction of the same number of samples using commercial kits would take 5-6 hours of work in a Biosafety Level 2 area with continuous use of a benchtop centrifuge. Additionally, PK treatment involves an estimated cost of 0.37 USD per sample whereas the cost of standard RNA extraction procedures is clearly superior (6-10 USD per sample in Argentina). We should also highlight that the HID method combined with the 2019-nCoV CDC Diagnostic Panel is an inexpensive option since it does not require extra reagents as PK+HID.

The simplicity and relatively short period of time required for the protocol and the full-availability and low price of the required reagents place PK+HID as a cheap, simple and fast alternative to traditional RNA extraction protocols.

## Data Availability

All data is included within the manuscript and supplementary information.

## Acknowledgments

V. L. thanks J. C. Reboreda and A. Quaglino for their support.

## Supporting information

**S1 Fig.**
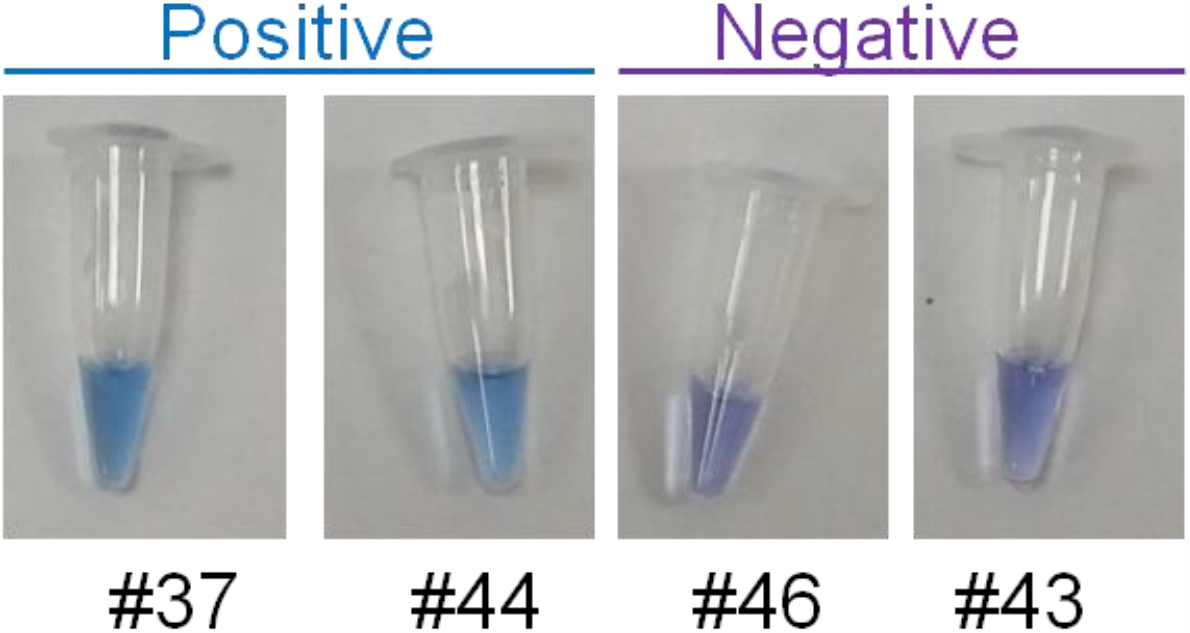
PK+HID can be combined with a detection kit based on loop-mediated isothermal amplification reactions. Representative colorimetric reactions run using COVID-19 Neokit Tecnoami, based on loop-mediated isothermal amplification. Diagnostics is made according to final color of the solution: blue, positive; violet, negative.

**S1 Table.**
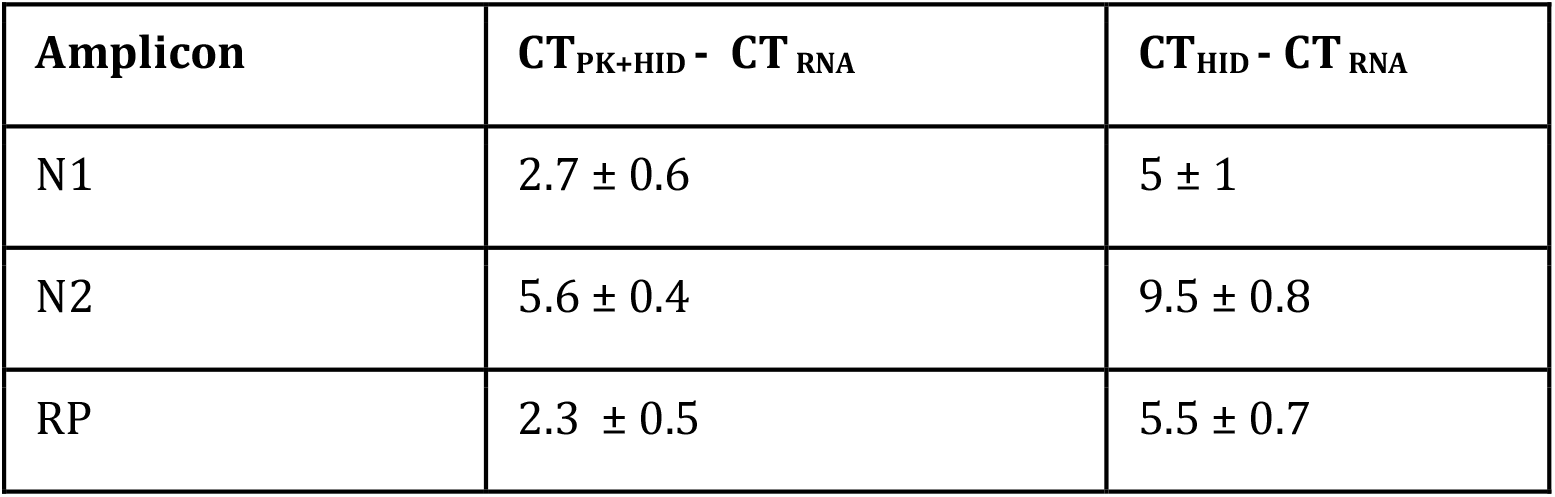
Mean variations in CT values in HID and PK+HID samples compared to purified RNA samples. Values are expressed as mean ± standard error.

**S2 Fig.**
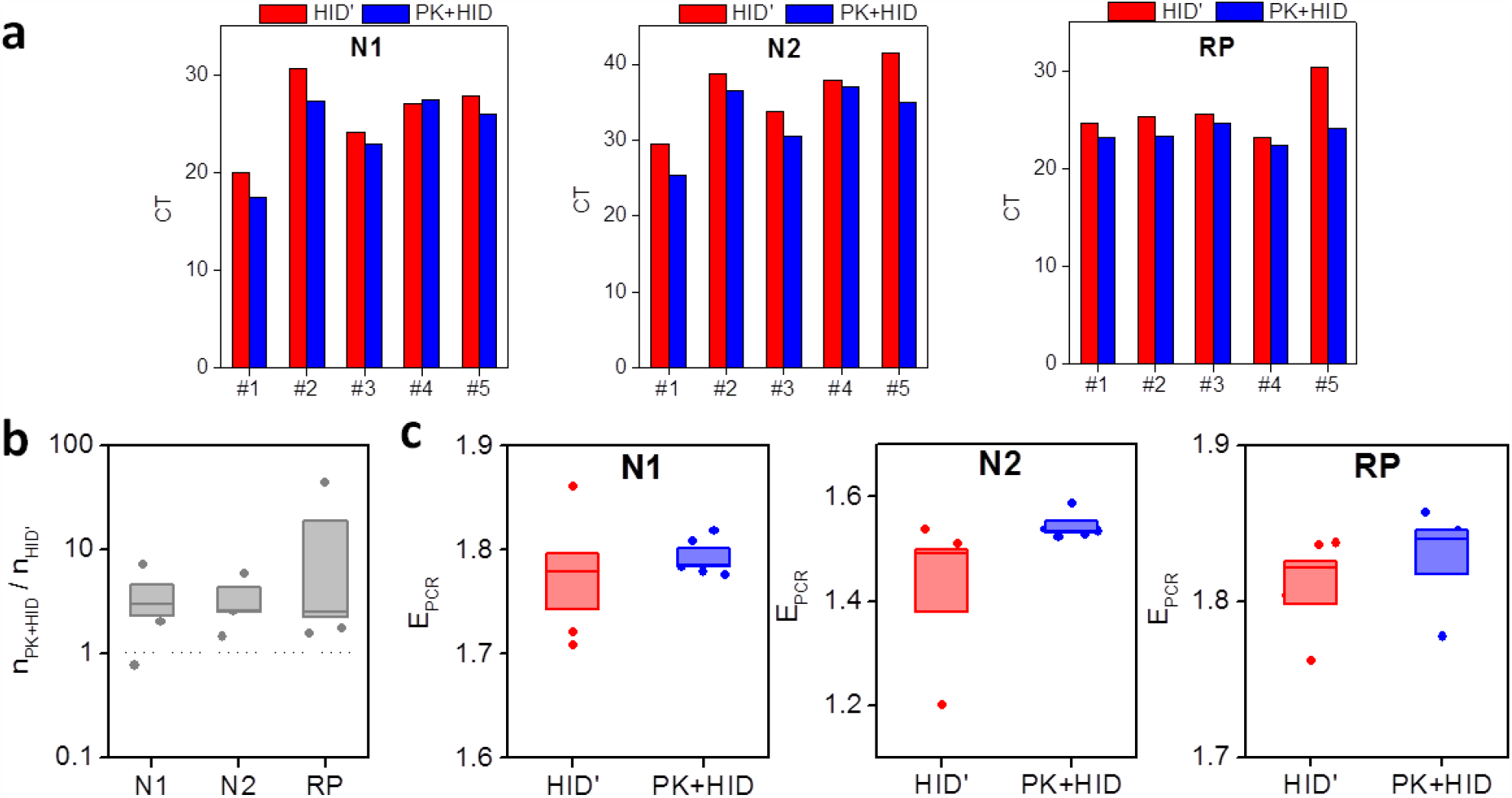
Effect of saline solution dilution in PK+HID method. Five positive nasopharyngeal swab samples (#1 to #5) were processed by adding 10 µl of proteinase K 10mg/ml (PK+HID samples) or 10 µl in proteinase K buffer (HID’ samples) and subjected to thermal incubations (55°C for 15 min and 98°C for 5 min). The viral N1 and N2 genes and the human RNase P gene (RP) were amplified and detected by RT-qPCR. **(a)** CT values obtained from RT-qPCR analysis of the same samples prepared by both different methods. **(b)** Ratio between relative amplicon amounts (n) of PK+HID and HID’ samples. The median of each measurement is represented with a line in the bars and the lengths of these bars represent the standard error. **(c)** Amplification efficiencies (E_PCR_). The median of each measurement is represented with a line in the bars and the lengths of these bars represent the standard error.

**S2 Table.** CT values obtained in RT-qPCR analysis of the same HID+PK samples introducing 5 or 10 µl of sample volume as input for RT-qPCR.

| CT<sub>5μL</sub> (gene) | CT<sub>10μL</sub> (gene) |
| --- | --- |
| 40.4 (N1) | 34.0 (N1) |
| 23.9 (RP) | 23.9 (RP) |
| 22.8 (N1) | 21.3 (N1) |
| 22.1(RP) | 21.4(RP) |
| 22.6 (N1) | 21.9 (N1) |
| 28.6 (RP) | 28.3 (RP) |
| 24.8 (N1) | 24.6 (N1) |
| 28.4 (RP) | 28.1 (RP) |
| 26.1 (N1) | 25.0 (N1) |
| 28.9 (RP) | 28.5 (RP) |
| 31.2 (N1) | 30.9 (N1) |
| 28.6 (RP) | 28.0(RP) |
| 29.6 (N1) | 28.9 (N1) |
| 30.0 (RP) | 29.5 (RP) |
| 34.1 (N1) | 35.6 (N1) |
| 29.5 (RP) | 29.0 (RP) |

**S3 Table.** CT values of false-negative samples detected with the PK+HID method, using the results obtained with purified RNA samples as reference.

| **CT<sub>RNA</sub> (gene, kit)** | **CT<sub>PK+HID</sub> (gene, kit)** |
| --- | --- |
| 30.7 (N, DisCoVery) | nd (N, DisCoVery) |
| 30.9 (ORF1ab, DisCoVery) | nd (ORF1ab, DisCoVery) |
| 27.4 (RP, DisCoVery) | 23.2 (RP, DisCoVery) |
| 23.5 (N, DisCoVery) | nd (N, DisCoVery) |
| 21.1 (ORF1ab, DisCoVery) | nd (ORF1ab, DisCoVery) |
| 26.2 (RP, DisCoVery) | 26.0 (RP, DisCoVery) |
| 31.3 (N, GeneFinder) | 40.1 (N, GeneFinder) |
| 31.3 (RdRp, GeneFinder) | nd (RdRp, GeneFinder) |
| 32.1 (E, GeneFinder) | nd (E, GeneFinder) |
| 27.3 (RP, Genefinder) | 30.3 (RP, Genefinder) |
| 34.6 (N, GeneFinder) | 41.3 (N, GeneFinder) |
| 34.5 (RdRp, GeneFinder) | nd (RdRp, GeneFinder) |
| 38.8 (E, GeneFinder) | nd (E, GeneFinder) |
| 27.2 (RP, Genefinder) | 28.2 (RP, Genefinder) |
| 35.9 (N, GeneFinder) | nd (N, GeneFinder) |
| 37.0 (RdRp, GeneFinder) | nd (RdRp, GeneFinder) |
| nd (E, GeneFinder) | nd (E, GeneFinder) |
| 28.4 (RP, Genefinder) | 30.6 (RP, Genefinder) |

**S3 Fig.**
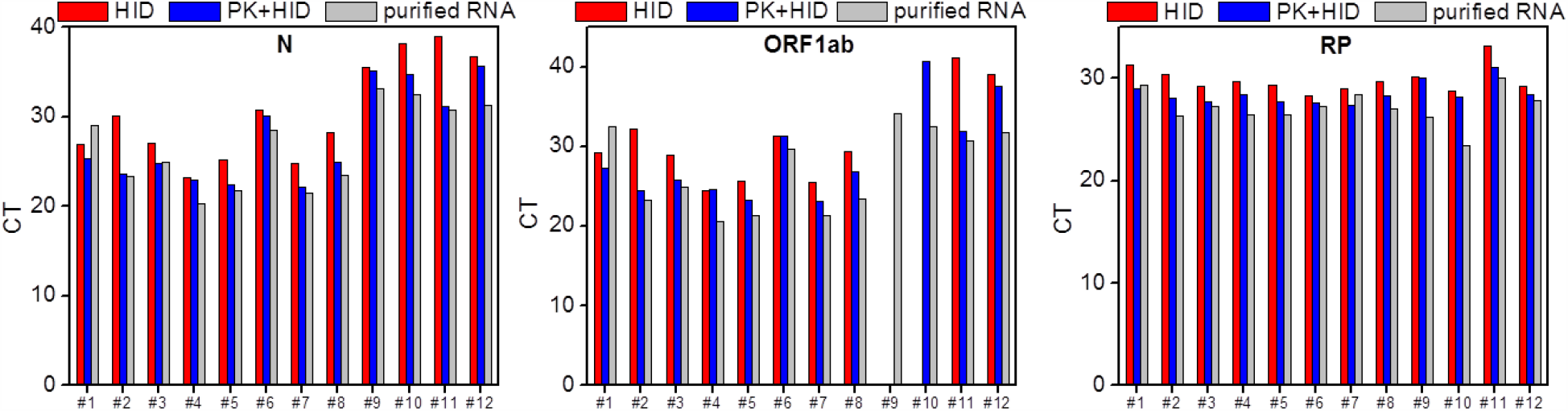
Performances of HID and PK+HID methods targeting N and ORF1ab amplicons. Twelve positive nasopharyngeal swab samples (#1 to #12) were processed by heat inactivation (HID samples), proteinase K treatment followed by heat inactivation (PK+HID samples) or RNA extraction (purified RNA samples). The viral N and ORF1ab genes and the human RNase P gene (RP) were amplified and detected by RT-qPCR. CT values obtained from RT-qPCR analysis of the same samples prepared by the three different methods.

**S4 Fig.**
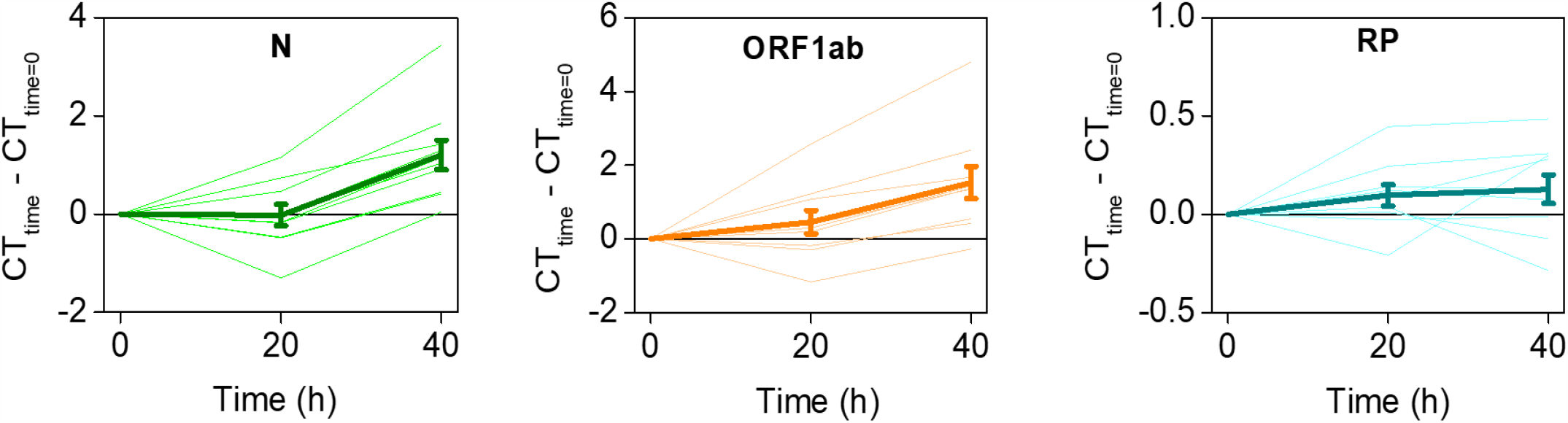
Stability of PK+HID samples at 4°C. Ten positive nasopharyngeal swab samples were processed by PK treatment followed by heat inactivation. These PK+HID samples were kept at 4°C. The viral N and ORF1ab genes and the human RNase P gene (RP) were amplified and detected by RT-qPCR at different times of incubation at 4°C (0, 20 and 40 h). Differences between CT values obtained at different times and time = 0 h. Thin, light lines represent individual sample measurements. Strong lines represent mean + SEM values.

**S5 Fig.**
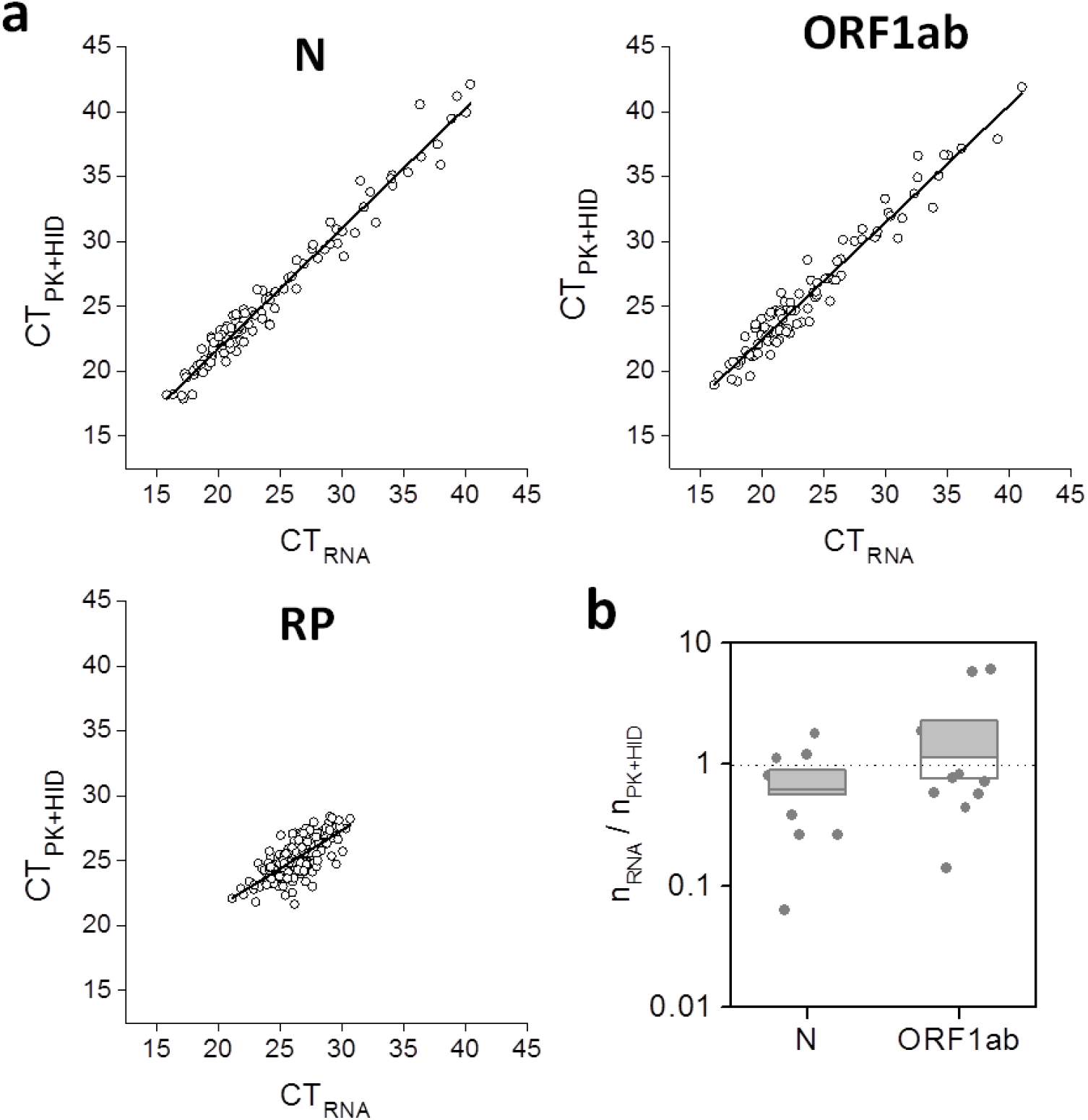
Performance of PK+HID method targeting N and ORF1ab amplicons. Randomly-selected nasopharyngeal swab samples were simultaneously processed for RNA extraction (purified RNA) or subjected to treatment with PK 1 mg/ml (55°C for 15 min) followed by heat-inactivation at 98°C for 5 min (PK+HID). The viral N and ORF1ab genes and the human RP gene were amplified and detected by RT-qPCR. **(a)** CT values obtained for the same samples prepared by the two different methods; the line obtained by regression of the data (continuous lines). RP plot also includes the data obtained in SARS-CoV-2 negative samples whereas N and ORF1ab plots include those data of negative samples that provided CT values above the positive-negative cut-off. **(b)** Ratio between the relative amplicon copy amounts detected using PK+HID or RNA purification for those nasopharyngeal swab samples that presented CT values >30 for any of the viral genes in purified RNA samples (N_data_ =10 and 12 for N and ORF1ab genes, respectively).

